# The ratio of triglycerides to total lipids in chylomicrons and extremely large VLDL, very large VLDL, large VLDL, medium VLDL, and small VLDL has a significant negative causal relationship with Coronary revascularization (ANGIO or CABG). A Mendelian randomization study

**DOI:** 10.1101/2025.04.28.25326612

**Authors:** Zhanqiang Yan, Jia Guo

## Abstract

**BACKGROUND:** Different lipids and lipoproteins have different relationships with atherosclerotic disease. We did Mendelian randomization study to explore the relationships between them.

**METHODS:** We conducted this Mendelian randomization (MR) study to analyze the causal relationship between TG levels/TG to total lipids ratio in lipoprotein subfractions and Coronary revascularization.

**RESULTS:** The ratio of TG to total lipids in chylomicrons(CM) and extremely large VLDL(P=2.43e-03, OR: 0.53, 95% CI: 0.36-0.80), very large VLDL(P=7.94e-04, OR: 0.68, 95%CI: 0.55-0.85), large VLDL(P=4.01e-06,OR:0.53,95%CI:0.41-0.69), medium VLDL(P=9.68e-03,OR:0.77,95%CI:0.63-0.94), small VLDL(P=0.02,OR:0.71,95%CI:0.53-0.94) showed a negative causal relationship with coronary revascularization, whereas this ratio of TG to total lipids in very small VLDL, IDL, large LDL, medium LDL, small LDL showed no causal correlation with it. After replacing the TG to total lipids ratio with TG levels the causal relationship became positive (except large VLDL). In contrast, when substituting the TG levels with TG-to-total-lipids ratio in very large HDL, large HDL, and medium HDL particles, their previously no causal relationship with coronary revascularization became positive.

Replacing TG levels with the TG-to-total-lipids ratio in in different sizes of VLDL particles enhances the particles’ anti-atherosclerotic effects while reducing pro-atherogenic activity. In contrast, the same replacement in HDL promotes pro-atherogenic effects and diminishes anti-atherogenic properties. This divergence is primarily attributed to their opposing impacts on Cholesteryl Ester Transfer Protein (CETP) function, which critically determines the net atherogenic outcome.

**CONCLUSIONS:** This ratio variation across lipoprotein types and sizes differentially modulates CETP activity. When CETP transfers cholesterol esters (CE) from HDL to other lipoproteins — such as chylomicrons (CM), VLDL, IDL, and LDL — these particles become substrates for a broader range of metabolism-related receptors.

Outcome 1: If this process enhances receptor binding, it reduces pro-atherogenic effects and may exert anti-atherosclerotic properties.

Outcome 2: Conversely, if impaired receptor binding occurs, the accumulation of unprocessed lipoproteins will inevitably promote atherosclerosis.

Different lipoproteins can bind to various receptors in tissues and cells, leading to either anti-atherosclerotic or pro-atherogenic effects. The final outcome depends on their net effect, which determines whether different lipoprotein subfractions exhibit a negative, positive, or neutral causal relationship with coronary revascularization.

## Introduction

During lipoproteins metabolism, as the lipids contained in them are continuously broken down for use by cells, the lipoproteins also bind to a wider variety of receptors.

Cholesteryl Ester Transfer Protein (CETP) is synthesized in the liver and in the plasma mediates the transfer of cholesterol esters from HDL to VLDL, chylomicrons, and LDL and the transfer of triglycerides from VLDL and chylomicrons to HDL[1]. Higher substrate concentrations increased CETP activity.

LPL is synthesized in muscle, heart, and adipose tissue. This enzyme hydrolyzes the TGs carried in chylomicrons and VLDL to fatty acids, which can be taken up by cells. The catabolism of triglycerides results in the conversion of chylomicrons into chylomicron remnants and VLDL into IDL (VLDL remnants). This enzyme requires Apo C-II as a cofactor. Apo A-V also plays a key role in the activation of this enzyme[1].

Lecithin: Cholesterol Acyltransferase (LCAT) is made in the liver. In the plasma, it catalyzes the synthesis of cholesterol esters (CE) in HDL by facilitating the transfer of a fatty acid from position 2 of lecithin to cholesterol[1].

CM and VLDL enriched in TG, CM contains apolipoproteins A-I, A-II, A-IV, A-V(promote LPL mediated TG lipolysis), B-48, C-II(Co-factor for LPL), C-III, and E(Ligand for LDL receptor), VLDL contains Apo B-100(Structural protein, Ligand for LDL receptor), C-I(Activates LCAT), C-II (Co-factor for LPL), C-III, and E(Ligand for LDL receptor). The larger the VLDL, the higher TG it contains, the more TG can be transported to HDL in exchange for receiving CE from HDL.

Different lipoproteins contain varying types and levels of lipids, and their ability to bind to a broad range of receptors in various tissues/cells differs. These differences lead to distinct anti-atherosclerotic or pro-atherogenic effects.

Since the early discovery of the LDL receptor, many other related receptors that bind to native or modified lipoproteins have been identified. In addition to the initially assigned role as cholesterol transporters, these LDL receptor-related proteins have a wide range of additional functions that influence diverse physiological and pathological processes, including embryonic development, inflammation, haemodynamics, thrombosis, neointima hyperplasia, and atherosclerosis[3][4][5][6][7][8]. The lipoprotein receptors are expressed in multiple types of vascular cells, and they mediate actions of a broad range of ligands to accelerate or block atherogenesis[3].

The LDLR family comprises a group of endocytic receptors on the cell surface, which bind and internalize extracellular ligands, including lipoproteins, exotoxins, and lipid-carrier complexes[9][10]. To date, numerous members of the LDLR family are reported that participate in a wide range of physiological processes[9][11]. In particular, LDLR, VLDLR, LRP5/6, LRP1, and LRP2 play a pivotal role in cholesterol homeostasis and lipid metabolism[9][12][13][14][15][16]. ApoE containing VLDL not only binds to VLDLR, but to LDLR, LRP8, LRP1, LRP2, and likely LRP6 as well[9] LDLR, LRP1, LRP 2, LRP5/6, ApoER2, and VLDLR belong to the LDL receptor family. Cluster of differentiation 36 (CD36 also referred as SR-B2), scavenger receptor class B type 1 (SR-B1), and lectin-like oxidized low-density lipoprotein receptor-1 (LOX-1) belong to a large family of pattern recognition receptors. They are expressed in multiple cell types including hepatocytes, vascular SMCs, neurons, macrophages, fibroblasts, and endothelial cells [3][17][18][19]. These receptors interact with a broad range of ligands including LDL, IDL, VLDL, oxLDL, circulating native or modified lipoproteins, to promote or inhibit atherogenesis. Here are some examples.

The LDL receptor regulates cholesterol homeostasis by the receptor-mediated endocytosis of LDL particles, and that the mutations in the LDL receptor gene cause elevated serum LDL cholesterol and coronary atherosclerosis in familial hypercholesterolaemia patients[3][20].

LRP5 is widely expressed in multiple tissues, including the liver, where the receptor is implicated in LDL and chylomicron clearance[3][21][22]. In sum, LRP5 deficiency and resulting downregulation of Wnt pathway contributes to exaggeration atherosclerosis, likely due to hypercholesterolaemia, systemic, or local inflammation and macrophage migration in the arterial wall[3].

In summary, current evidence indicates a dichotomous role of endothelial SR-BI in atherogenesis. The receptor mediates anti-inflammatory effects of HDL, contributing to increased bioavailable NO and protection against atherosclerosis. On the other hand, in conditions of high plasma LDL, SR-BI facilitates LDL transport across the endothelial layer into the artery wall, which leads to foam cell formation and progression of atherosclerosis[3].

The binding of different lipoproteins to different receptors will produce different anti-atherosclerotic or pro-atherogenic effects.

The LDL receptor is present in the liver and most other tissues. It recognizes Apo B-100 and Apo E and hence mediates the uptake of LDL (LDL is a product of VLDL metabolism), chylomicron remnants, and IDL (VLDL remnants enriched in cholesterol esters), and they can be removed from the circulation by the liver via binding of Apo E to LDL and LRP and other hepatic receptors[1].

These lipoprotein particles (VLDL, IDL, and LDL) are rich in cholesteryl esters (CEs), which-unlike CEs in HDL-are not transferred to other lipoproteins. (In HDL, CEs can be transferred to VLDL, chylomicrons, and LDL if the load is excessive.)

Their metabolic fate involves two potential pathways:

1. “Metabolism-related anti-atherogenic pathway”: Clearance via receptors (e.g., LDL receptor, LRP1, VLDL receptor).
2. “Non-metabolism-related pro-atherogenic pathway”: Prolonged circulation time, oxidation, intimal penetration, and uptake by macrophages, activating inflammatory cells (e.g., endothelial cells, monocytes, T cells).

CETP transfers CE from HDL to various lipoproteins (e.g., CM, VLDL, IDL, LDL), which are then recognized and processed by diverse receptors. Lipoproteins vary in composition (e.g., lipid levels and ratios), and their receptors are differentially expressed across tissues and cells. These interactions determine whether lipoproteins exert anti-atherogenic or pro-atherogenic effects. (Figure 1)

**Figure 1:**
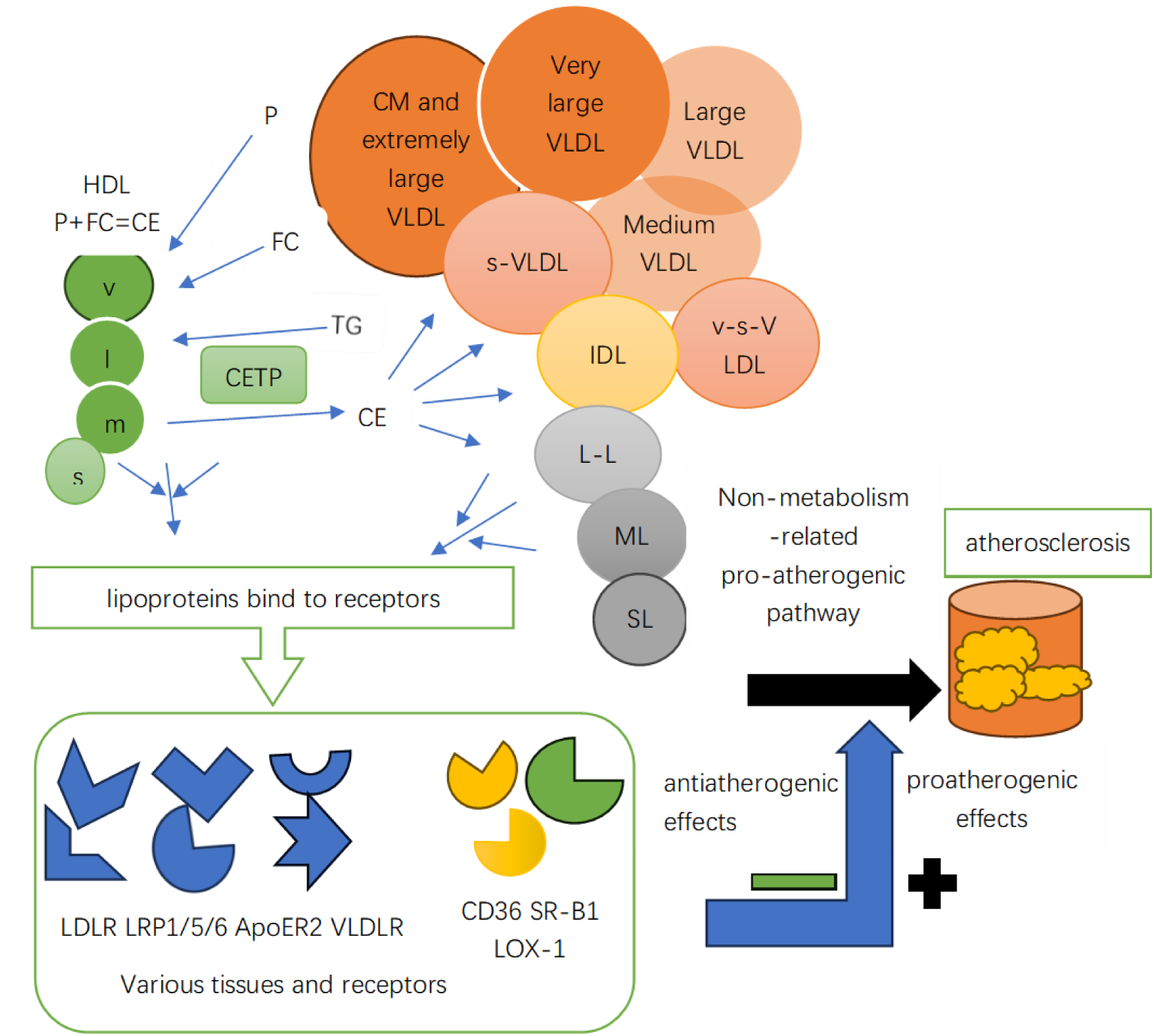
Lipoproteins and Receptors P:phospholipids,FC:free cholesterol,CE;cholesterol esters,C;cholesterol· TG: triglycerides, CETP: Cholesteryl Ester Transfer Protein, HDL;Hígh-Density Lipoproteins, V:very large HDL, Llarge HDL, M;medium HDL, S;small HDL, CM;chylomicrons, VLDL·Very Low-Density Lipoproteins, IDL·lntermediate-Density Lipoproteins, LDL;Low-Density Lipoproteins, s-VLDL:small VLDL, v-s-VLDL:very small VLDL, L-L·large LDL, ML;medium LDL, SL·small LDL, LDLR;LDL receptor, LRPl/5/6:low-density lipoprotein receptor-related proteinl/5/6, ApoER2:apolipoproteiπ E receptor 2, VLDLR;VLDL receptor, CD36: cluster of differentiation 36, SR-B1: Class B Scavenger Receptor BI, LOX-1: lectin-like oxidized low-density lipoprotein receptor-1.

Mendelian randomization is an application of instrumental variable analysis, which aims to test a causal hypothesis in non-experimental data. In an MR analysis, genetic variants, commonly single nucleotide polymorphisms (SNPs), are used as instrumental variables (IVs) for the putative risk factor[2]. The transmission of DNA from parents to offspring follows Mendel’s second law. It can achieve the purpose similar to the randomization in randomized controlled trials, equivalent to natural randomization, can achieve the purpose of reducing the influence of confounding factors, and avoid the effect of reverse causation. Large-scale genome-wide association studies (GWAS) in humans have revealed numerous genetic variants, which can be used as instrumental variables (IVs) to conduct the Mendelian randomization (MR) studies. More and more genetic variants associated with coronary heart disease, lipid traits and other disease traits have been uncovered by large-scale genome-wide association studies (GWAS), more and more Mendelian randomization studies will be applied to a wider range of fields. Mendelian randomization has been increasingly applied over recent years to predict the efficacy and safety of existing and novel drugs targeting cardiovascular risk factors and to explore the repurposing potential of available drugs[2].

## Methods

### Design

Applying a two-sample MR framework, we analyzed instrument-exposure and instrument-outcome associations derived from independent GWAS summary statistics to infer causal relationships between [exposure] and [outcome], implementing inverse-variance weighted regression as the primary analytical method. Single nucleotide polymorphisms (SNPs) are used as instrumental variables (IVs).The genetic variant (or multiple genetic variants) used as instrumental variable(IVs) for the risk factor must (i) reliably associate with the risk factor under investigation (relevance assumption); (ii) not associate with any known or unknown confounding factors (independence assumption); and (iii) influence the outcome only through the risk factor and not through any direct causal pathway (exclusion restriction assumption)[2]. (Figure 2)

**Figure 2:**
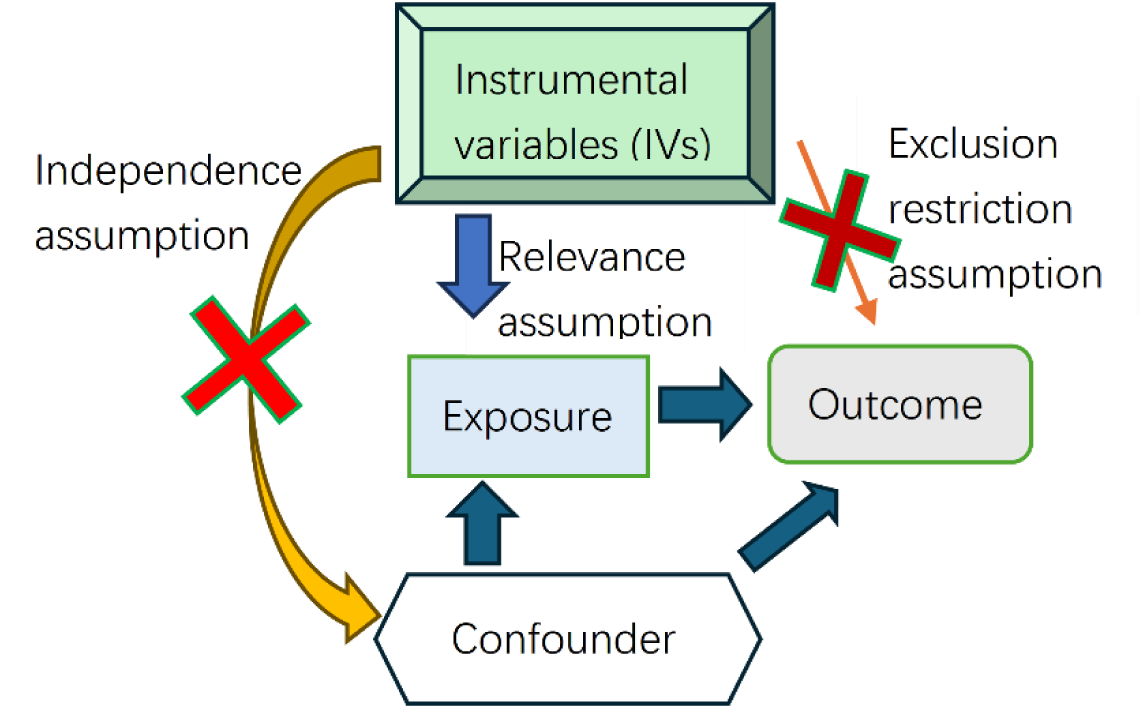
Graphical Abstract Exposure: Triglycerides to total lipids ratio in lipid sub-fractions Triglyceride levels in lipid sub-fractions Outcome: Coronary revascularization (ANGIO or CABG)

### Data

The outcome data was sourced from FinnGen, while the exposure data was obtained from the IEU OpenGWAS project (https://gwas.mrcieu.ac.uk/datasets/), ensuring no population overlap between the two samples. All study participants were of European descent.

Table 1 show the detailed information about the datasets. All the information about the exposure data sets is the same as triglyceride levels in LDL differing only in GWAS-ID. The GWAS-ID of other exposure data sets can be seen in Table 3. So only three exposure data sets are listed in Table 1. Since all data used was published previously in the public database, no additional ethical approval was needed.

**Table 1.**
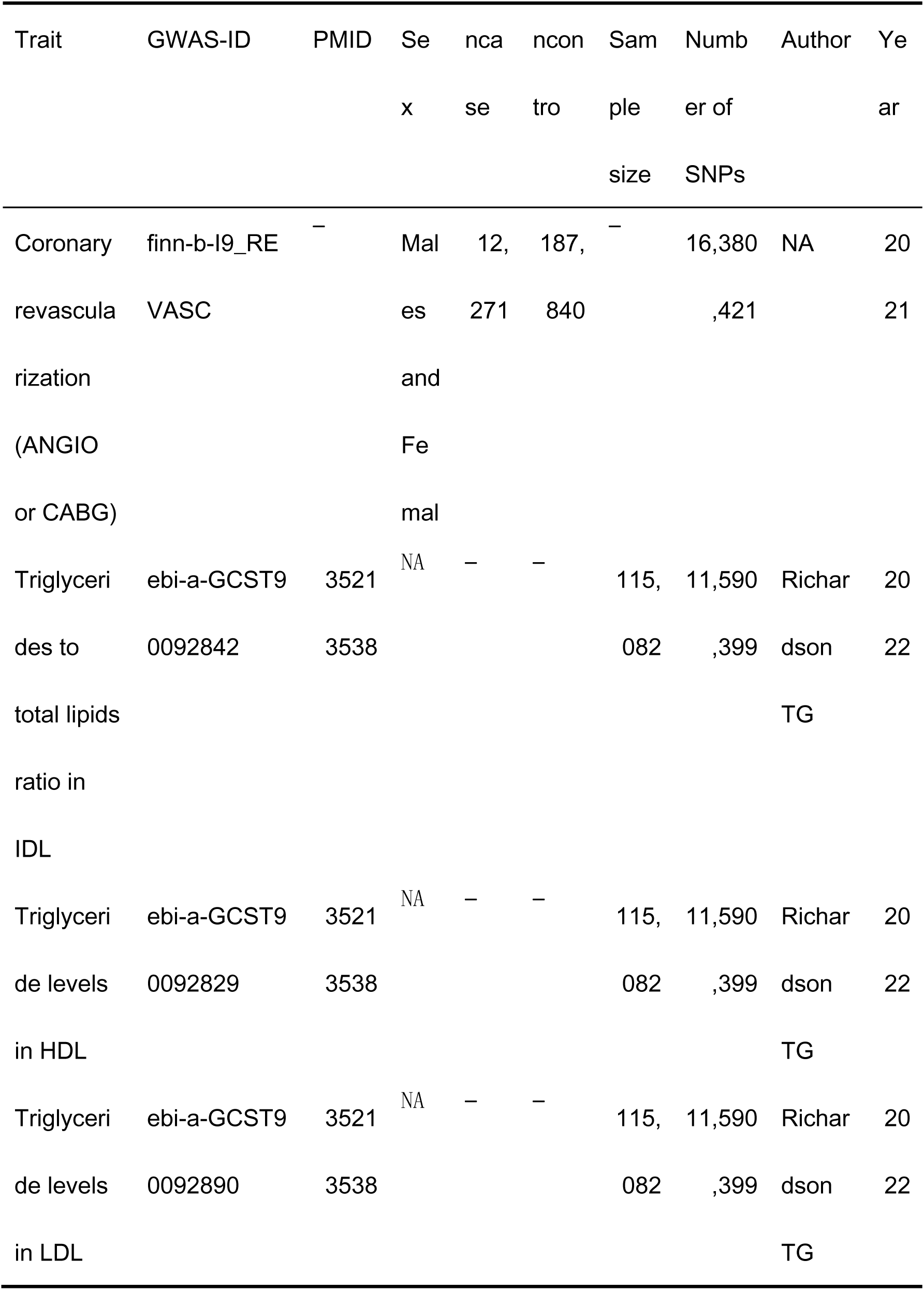
Characteristics of data of Exposures/Outcomes. Population: European.

We use the “extract_instruments” function in R package“TwoSampleMR” to extract instrumental variables (IVs), the SNPs selected as instrumental variables (IVs) highly correlated with exposure traits (p < 5 × 10^-8^). In order to avoid the linkage disequilibrium, all SNPs are clumped under a strict clump window (r^2^ = 0.001 and kb = 10,000). We calculated the F-statistic, all SNPs (with an F-statistic greater than ten) can be considered highly effective in mitigating potential bias.

### Statistical Analyses

In a two-sample MR study based on multiple genetic variants and summarized data, the causal estimate can be obtained by the inverse-variance weighted method, which is a meta-analysis of the single Wald ratios and is the most efficient method (greatest statistical power)[2]. The “ TwoSampleMR ” in R software(version 4.4.1) and “MR-PRESSO” packages were used to conduct this MR analysis. We used inverse variance weighted (IVW), MR Egger, weighted median, Simple mode, and weighted mode five MR analytical methods. The “harmonise_data” function in the “TwoSampleMR” package harmonizes the effect alleles and effect sizes across datasets. We used Cochran ’ s Q statistic to test for heterogeneity. If the inverse-variance weighted (IVW) Cochran ’ s Q test yielded p< 0.05, indicating significant heterogeneity, we applied the random-effects IVW method.

To assess horizontal pleiotropy, we employed the “mr_pleiotropy_test” function in “TwoSampleMR”. A significant result (p< 0.05) suggested the presence of horizontal pleiotropy, prompting us to perform an “MR-PRESSO” test to identify and remove outlier SNPs, thereby correcting the results.

## Results

### The MR Results

Figure 3 shows the results of IVW method. The random-effect IVW method was the primary analysis. The Mendelian randomization (MR) study results of the other four methods can be seen in supplemental material.

**Figure 3:**
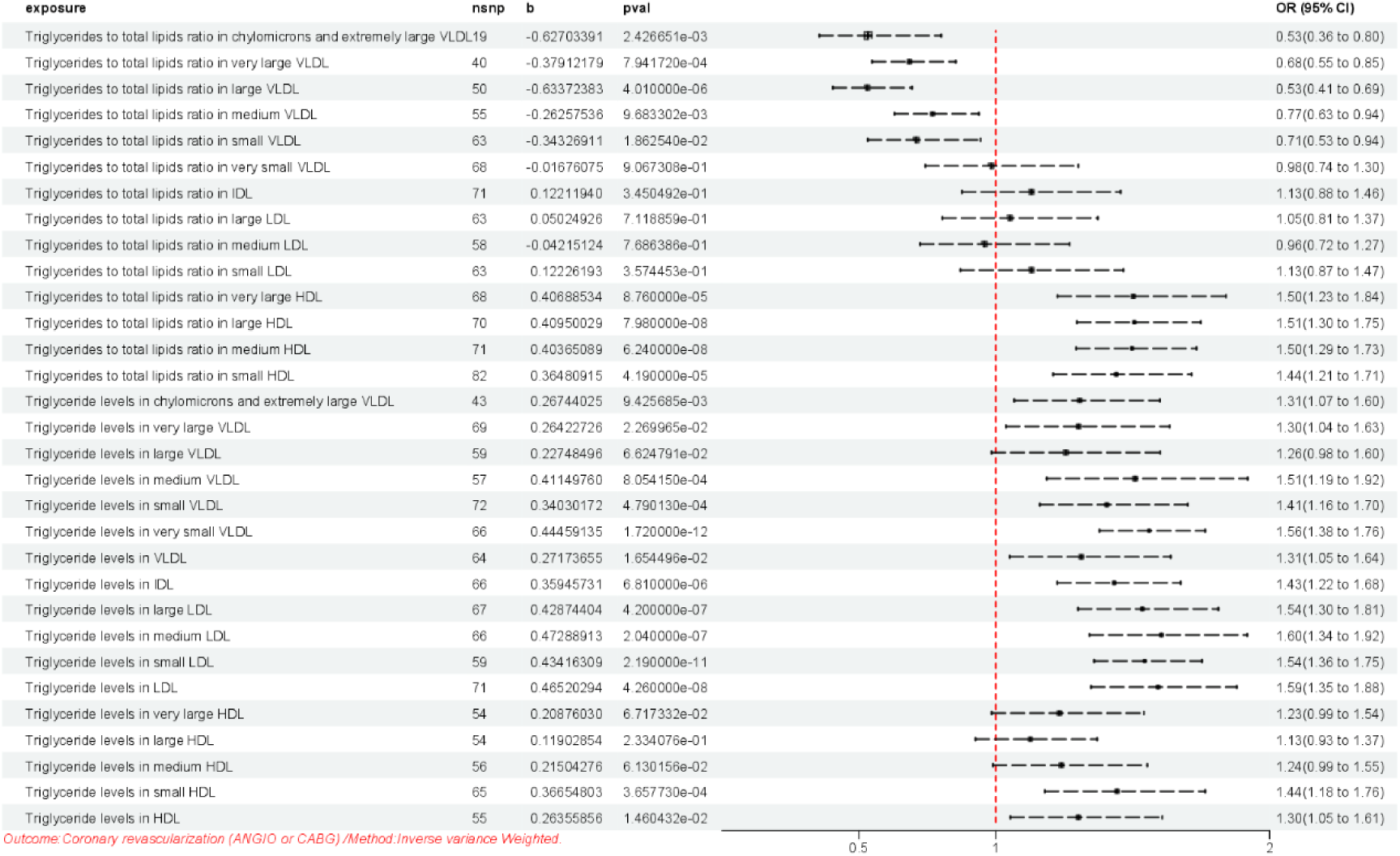
The Mendelian randomization (MR) study results. Outcome: Coronary Revascularization/Method: inverse variance weighted (IVW)

The causal relationship between lipoprotein subfractions and coronary revascularization demonstrates distinct patterns based on triglyceride (TG) composition and particle characteristics. Specifically:

1. Inverse causal relationship was found in:

*TG to total lipids ratio in:

CM and extremely large VLDL (P=2.43e-03, OR: 0.53, 95% CI: 0.36-0.80)

Very large VLDL (P=7.94e-04, OR: 0.68, 95%CI: 0.55-0.85)

Large VLDL (P=4.01e-06, OR:0.53,95%CI:0.41-0.69)

Medium VLDL (P=9.68e-03, OR:0.77,95%CI:0.63-0.94)

Small VLDL (P=0.02, OR:0.71,95%CI:0.53-0.94)

2. Null causal relationship was observed in:

*TG to total lipids ratio in:

Very small VLDL (P=0.91, OR:0.98,95%CI:0.74-1.30)

IDL (P=0.35, OR:1.13, 95%CI:0.88-1.46)

Large LDL (P=0.71, OR:1.05, 95%CI:0.81-1.37)

Medium LDL (P=0.77, OR:0.96, 95%CI:0.72-1.3)

Small LDL (P=0.36, OR:1.13, 95%CI:0.87-1.47)

*TG levels in:

Large VLDL (P=0.07, OR:1.26,95%CI:0.98-1.60)

Very large HDL (P=0.07, OR:1.23,95%CI:0.99-1.54)

Large HDL (P=0.23, OR:1.13, 95%CI:0.93-1.37)

Medium HDL (P=0.06, OR:1.24, 95%CI:0.99-1.55)

3. Positive correlation emerged in:

*TG to total lipids ratio in:

Very large HDL (P=8.76E-05, OR:1.50,95% CI:1.23-1.84)

Large HDL (P=7.98E-08, OR:1.51,95%CI:1.30-1.75)

Medium HDL (P=6.24E-08, OR:1.50,95%CI:1.29-1.73)

Small HDL (P=4.19E-05, OR:1.44,95% CI:1.21-1.71)

*TG levels in:

CM and extremely large VLDL (P=9.42E-03, OR:1.31,95%CI:1.07-1.60)

Very large VLDL (P=0.02, OR:1.30,95%CI:1.04-1.63)

Medium VLDL (P=8.05 E-04, OR:1.51,95%CI:1.19-1.92)

Small VLDL (P=4.79E-04, OR:1.41,95% CI:1.16-1.70)

Very small VLDL (P=1.72E-12, OR:1.56,95%CI:1.38-1.76)

VLDL (P=0.02, OR:1.31,95% CI:1.05-1.64)

IDL (P=6.81E-06, OR:1.43,95% CI:1.22-1.68)

Large LDL (P=4.20E-07, OR:1.54,95%CI:1.30-1.81)

Medium LDL (P=2.04E-07, OR:1.60,95% CI:1.34-1.92)

Small LDL (P=2.19E-11, OR:1.54,95%CI:1.36-1.75)

LDL (P=4.26E-08, OR:1.59 ,95% CI:1.35-1.88)

Small HDL (P=3.66E-04, OR:1.44,95% CI:1.18-1.76)

HDL (P=0.01, OR:1.30,95% CI:1.05-1.61).

### Sensitivity Analyses

To assess horizontal pleiotropy, we employed the “mr_pleiotropy_test” function in “TwoSampleMR”. A significant result (p<0.05) suggested the presence of horizontal pleiotropy in triglycerides to total lipids ratio in(very large VLDL[P=0.019455006], medium VLDL [P=0.040858129]), and triglyceride levels in (chylomicrons and extremely large VLDL [P=0.027157549], very small VLDL [P=0.012959631], small LDL[P=0.041125941]). There was no horizontal pleiotropy (p > 0.05) in the others.

Tble2 shows outliers reported by the “MR-PRESSO” test. The table lists the respective GWAS-ID numbers and outlier SNPs.

**Tble2:** Outlier SNPs reported by the “MR-PRESSO” test.

| expo<br>sure | Triglycerides<br>to total lipids<br>ratio in very<br>large VLDL \|\|<br>id:ebi-a-GCS<br>T90093027 | Triglycerides<br>to total lipids<br>ratio in<br>medium<br>VLDL \|\|<br>id:ebi-a-GCS<br>T90092927 | Triglyceride<br>levels in<br>chylomicrons<br>and<br>extremely<br>large VLDL \|\|<br>id:ebi-a-GCS<br>T90093050 | Triglyceride<br>levels in very<br>small VLDL \|\|<br>id:ebi-a-GCS<br>T90093038 | Triglyceride<br>levels in<br>small LDL \|\|<br>id:ebi-a-GCS<br>T90092966 |
| --- | --- | --- | --- | --- | --- |
| out1 | rs10455872 | rs10455872 | rs10455872 | rs11231694 | rs10455872 |
| ier | rs11591147 | rs10750766 | rs116843064 | rs116843064 | rs116843064 |
| SNPs | rs117733303 | rs11122450 | rs117733303 | rs17242381 | rs117733303 |
|  | rs12151108 | rs11591147 | rs12179053 | rs186696265 | rs1561748 |
|  | rs1260326 | rs117733303 | rs1561748 | rs261290 | rs17242381 |
|  | rs12679834 | rs12151108 | rs28601761 | rs2943652 | rs2972140 |
|  | rs12740374 | rs12740374 | rs2943635 | rs602633 | rs602633 |
|  | rs1532085 | rs2925979 | rs3127580 |  |  |
|  | rs174537 | rs2972143 | rs328 |  |  |
|  | rs1885659 | rs3127580 | rs895893 |  |  |
|  | rs2302263 | rs328 | rs964184 |  |  |
|  | rs28807203 | rs4336630 |  |  |  |
|  | rs2925979 | rs653178 |  |  |  |
|  | rs3127580 | rs693668 |  |  |  |
|  | rs3129774 | rs72836561 |  |  |  |
|  | rs4336630 | rs78058190 |  |  |  |
|  | rs62188784 | rs964184 |  |  |  |
|  | rs693668 | rs6905288 |  |  |  |
|  | rs72836561 | rs1561748 |  |  |  |
|  | rs964184 | rs1260326 |  |  |  |

Table3 shows the “mr_pleiotropy_test” results, after removing outlier SNPs. There was no horizontal pleiotropy (p > 0.05).

**Table3:**
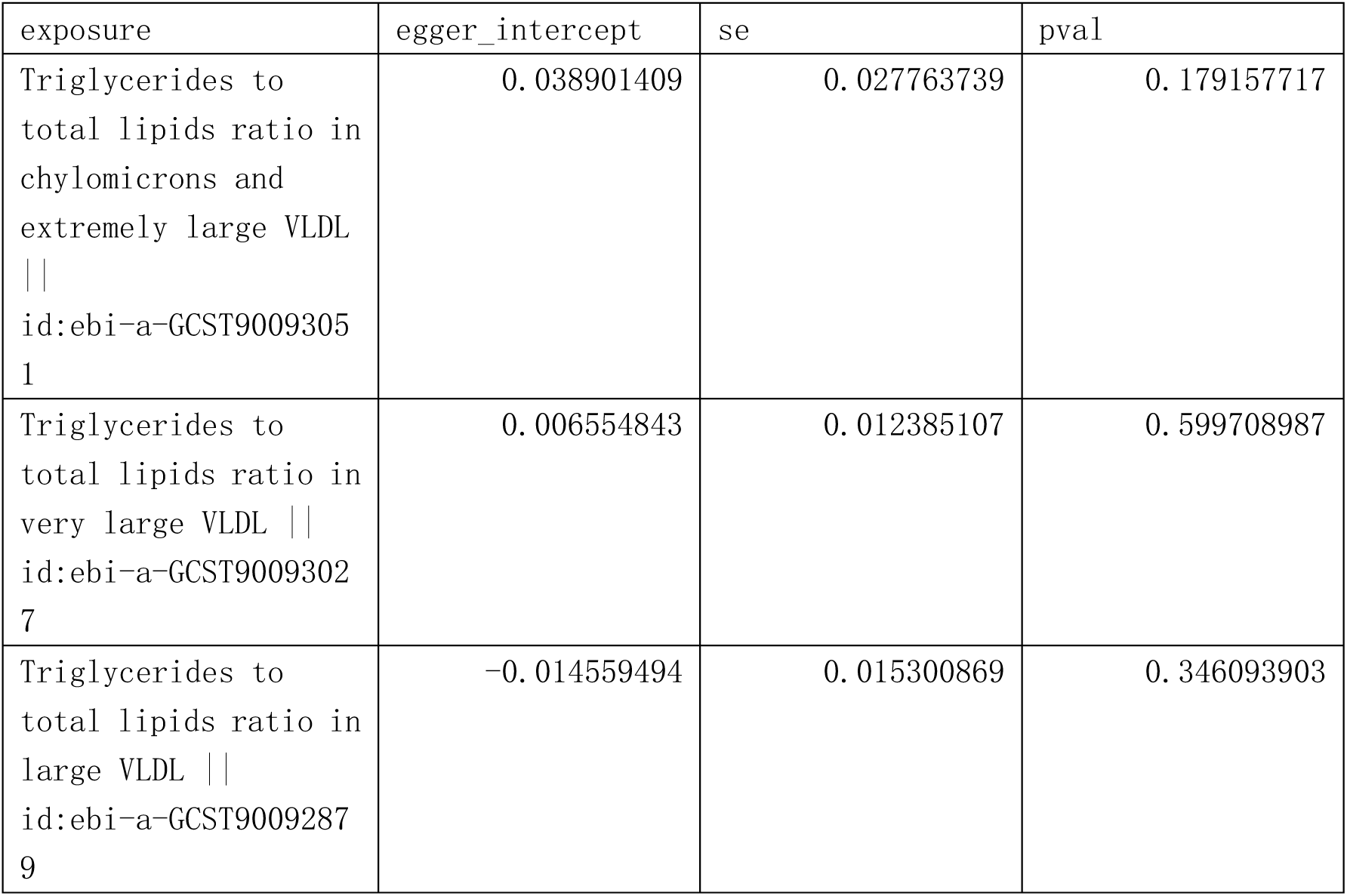
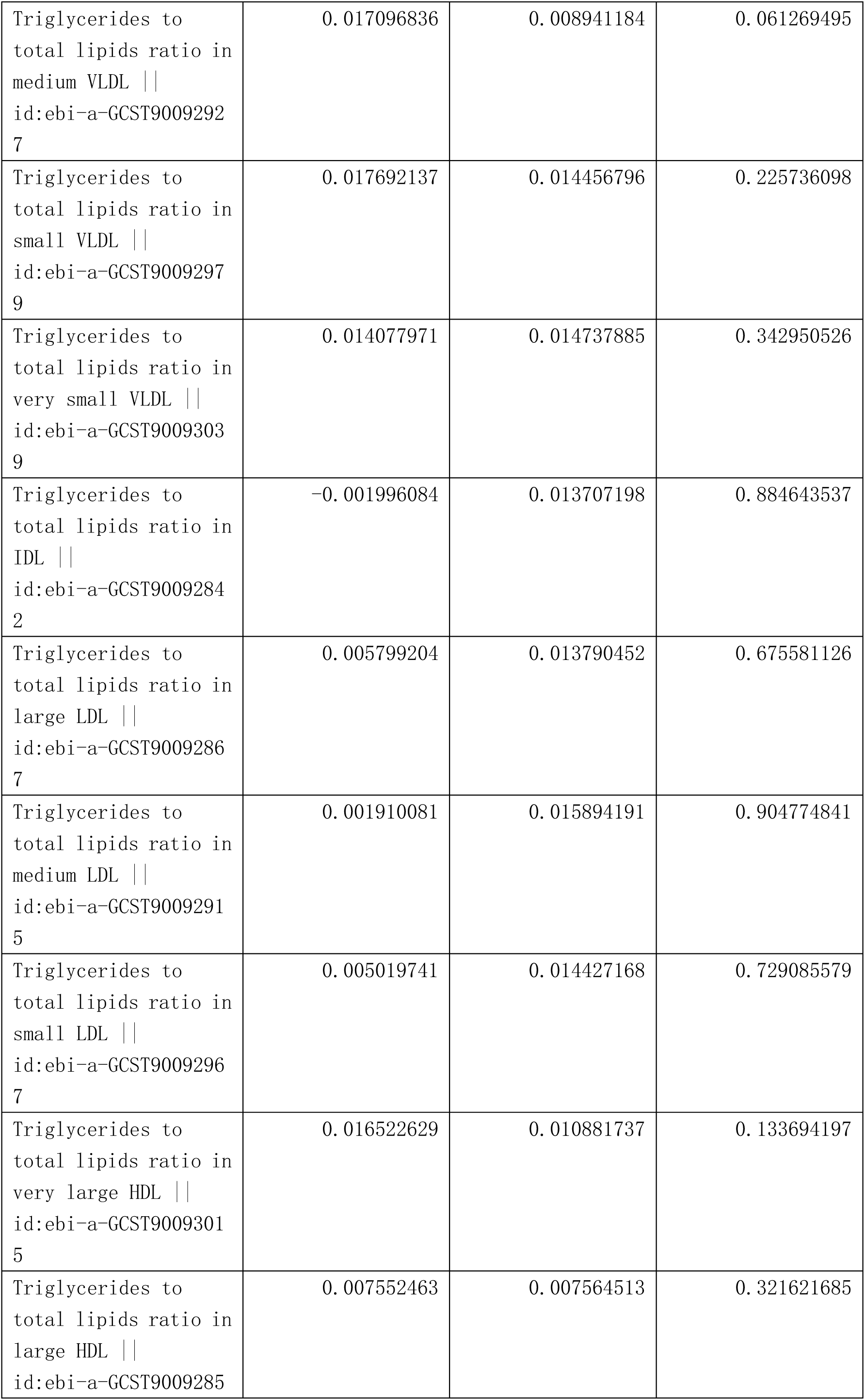
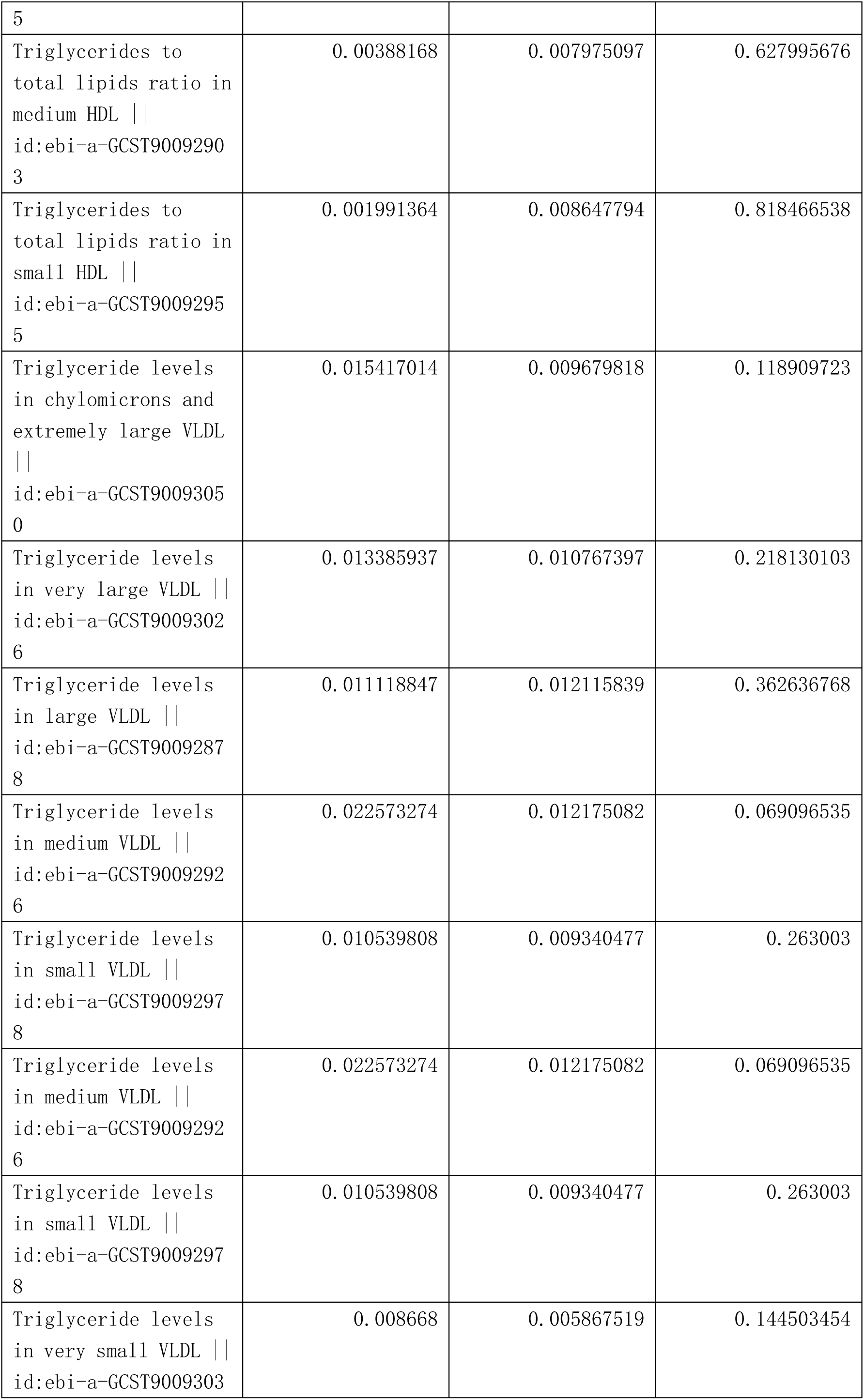
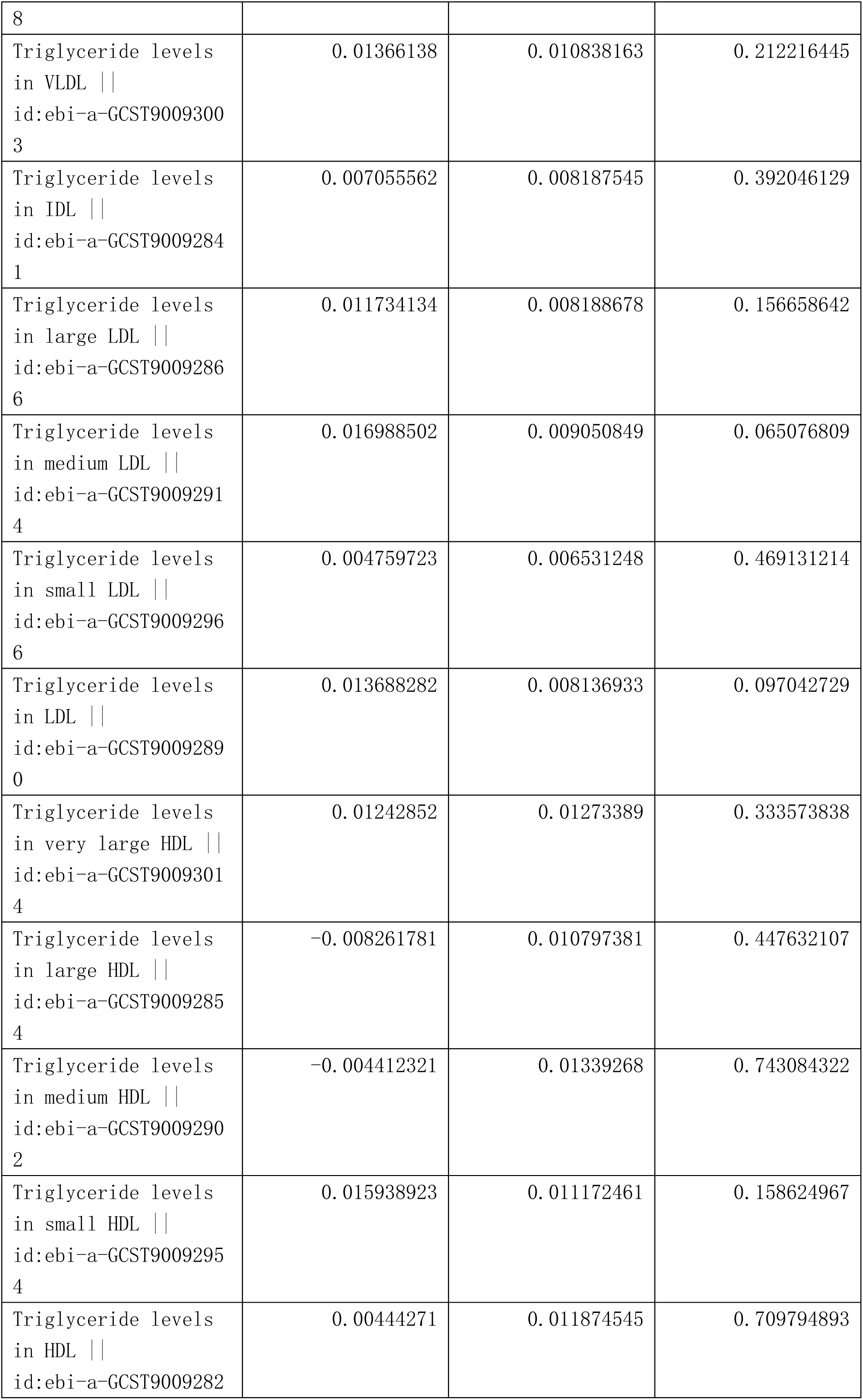
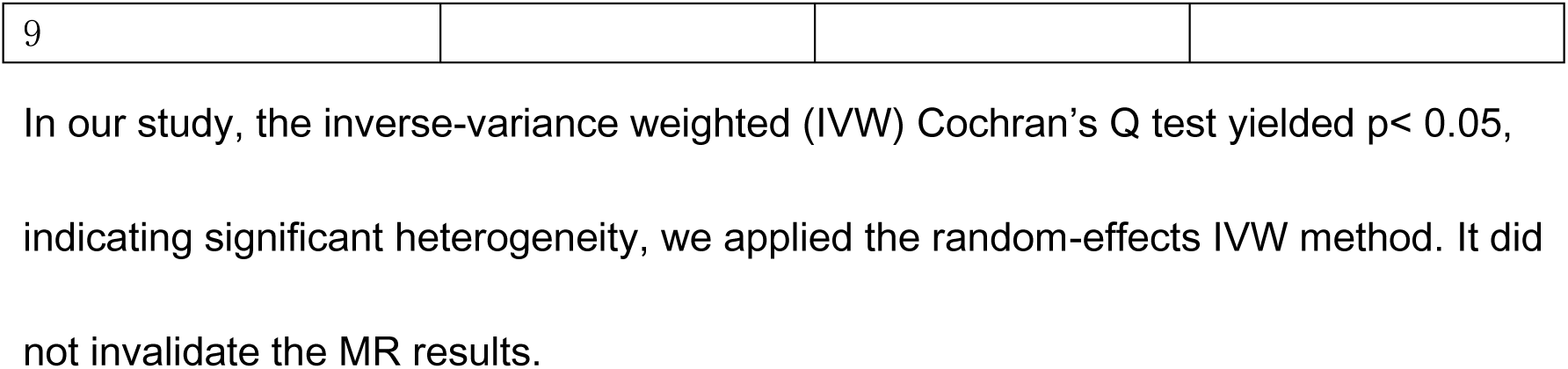
The “mr_pleiotropy_test” results, after removing outlier SNPs.

In our study, the inverse-variance weighted (IVW) Cochran’s Q test yielded p< 0.05, indicating significant heterogeneity, we applied the random-effects IVW method. It did not invalidate the MR results.

## Discussion

Our study reveals that the ratio of TG to total lipids in chylomicrons and extremely large VLDL, very large VLDL, large VLDL, medium VLDL, and small VLDL has a significant negative causal relationship with Coronary revascularization (ANGIO or CABG). The higher ratio of TGs to total lipids in VLDL can be higher levels of TG and lower CE levels. Both of them promote CETP to transfer CE from HDL to VLDL in exchange for TG to HDL. LCAT can catalyze a fatty acid from phospholipids to FC resulting in the formation of CE in HDL, promoting CE to move to the core of the HDL, allowing additional FC to be transferred to HDL[1]. CETP transferred CE from HDL to VLDL in exchange for TG to HDL can also promote this process. Newly formed HDL can also obtain cholesterol and phospholipids from chylomicrons and VLDL during their lipolysis by LPL[1]. Therefore, from the formation of CM and VLDL, the concentration of TG, phospholipids and FC in them is progressively decreases, while the concentration of CE keeps rising. So higher ratio of TGs to total lipids in CM and VLDL can not only promote the uptake of cholesterol by HDL and the synthesis of CE, but also promote the transport of CE from HDL to a wider variety of lipoproteins. It is beneficial to the reverse transport of cholesterol. Because cholesterol is transferred to a wider variety of lipoproteins, and can be processed by a wider variety of receptors.

Different types of lipoproteins bind to different receptors and have different metabolic processes in the liver and other tissues. It expands the liver’s ability to take up, process and expel cholesterol. Our study reveals that the higher ratio of TG to total lipids in CM and extremely large VLDL, very large VLDL, large VLDL, medium VLDL, and small VLDL promotes these processes, and has the following beneficial effects:1) promoting the uptake of cholesterol by HDL, 2) promoting the synthesis of CE, 3) promoting the transport of CE from HDL to a wider variety of lipoproteins (such as CM,VLDL,IDL,LDL), 4) cholesterol can be recognized and processed by a wider range of receptors, such as LDL receptors, LDL Receptor Related Protein 1(LRP-1), VLDL receptors and other hepatic receptors, expanding the clearance pathway of cholesterol, 5)expanding the liver’s ability to uptake, process and expel cholesterol, 6) increasing the clearance of cholesterol through metabolism-related receptor pathways for more cellular use, and reduce “ non-metabolism-related pro-atherogenic pathways ” , 7) These different metabolic processes affect cholesterol reverse transport in a wider range of interactions, 8) These different metabolic processes affect other types of lipids (such as TG, FC and phospholipids) and lipoprotein metabolism in a wider range of interactions. Which we could temporarily name as “promote-cholesterol-metabolism-effects”. Our study reveals that higher ratio of TGs to total lipids in CM and extremely large VLDL, very large VLDL, large VLDL, medium VLDL, and small VLDL promotes CETP to transfer CE from HDL to VLDL in exchange for TG to HDL, promotes the reverse cholesterol transport, promotes lipoproteins to bind to corresponding metabolism-related receptors, and reduces the “ non-metabolism-related pro-atherogenic pathway ” , produces anti-atherosclerotic effects. And there is a significant negative causal relationship with Coronary revascularization.

Our study reveals that the causal relationship between TG to total lipids ratio in VLDL sub-fractions and Coronary revascularization shifts from negative (TG to total lipids ratio in chylomicrons and extremely large VLDL, very large VLDL, large VLDL, medium VLDL, small VLDL has a significant negative causal relationship with Coronary revascularization) to no (TG to total lipids ratio in very small VLDL, IDL, large LDL, medium LDL, small LDL has no causal correlation with Coronary revascularization). The difference between them is that as the metabolism progresses, their lipid levels changed. From the beginning of the formation of CM and VLDL, the concentration of TG contained in it is constantly decreasing. And CE levels are constantly rising. Higher concentration of TG in VLDL promotes the exchange (CETP transferring CE from HDL to VLDL in exchange for TG to HDL), while higher concentration of CE in VLDL inhibits it. This state of affairs plays a beneficial role in the right degree and a disadvantageous role in the excessive case. For very small VLDL,IDL, large LDL, medium LDL and small LDL, these lipoproteins transport phospholipids and FC to HDL and receive CE from HDL, the “promote-cholesterol-metabolism-effects” can go from big to small. As metabolism progresses, smaller VLDL contain higher CE. The “harmful effects” (Elevated levels of cholesteryl ester (CE)-rich lipoproteins disrupt ligand-receptor interactions, particularly with LDL receptors, resulting in prolonged circulatory retention and enhanced endothelial deposition of atherogenic lipids.) grow from small to large. The causal relationship shifts from negative to no. Different types of lipoproteins bind to different receptors in various tissues and cells, and have different anti-atherogenic or pro-atherogenic effects.

Or another reason, For those smaller than small VLDL, because VLDL metabolize from large granules to small ones, with sufficient LPL to hydrolyze triglycerides and sufficient CETP to transfer CE from HDL to VLDL in exchange for TG to HDL, there is still a surplus of TGs, which means that too many cholesterol-rich particles were produced, exceeding the processing capacity of the receptor. It adds to the “non-metabolism-related pro-atherogenic pathway”. The causal relationship between TGs to total lipids ratio in these lipoproteins and Coronary revascularization changes from negative to no.

Our study reveals that the causal relationship between TG levels in VLDL sub-fractions and Coronary revascularization changes from positive to no (TG levels in CM, extremely large VLDL and very large VLDL have positive causal relationships with Coronary revascularization, while TG levels in large VLDL have no causal correlation with it), and then to positive(TG levels in medium VLDL, small VLDL, very small VLDL,IDL, large LDL, medium LDL, small LDL, and LDL have positive causal relationship with it). The causal relationship had turned, and this trend suggests that their deleterious effects in promoting atherosclerosis are gradually diminishing until they are none, and then turn from none to large. As metabolism proceeds, the “promote-cholesterol-metabolism-effect” goes from small(begin) to large(peak), and then from large to small. “Harmful effects” (The accumulation of CE-enriched lipoprotein particles impairs receptor-mediated clearance, promoting atherosclerotic plaque formation through reduced cellular uptake.) gradually decreased and then increased. In larger than large VLDL particles, Too much production of cholesterol-rich particles impaired receptor binding, and increasing the “non-metabolism-related pro-atherogenic pathway”. For those particles smaller than large VLDL, because VLDL metabolize from large granules to small ones, if there is still a surplus of TG, it means that too many cholesterol-rich particles are being produced. Excessive CE loading alters lipoprotein surface topology, sterically hindering apolipoprotein B-100 recognition by LDL receptors, thereby establishing a cholesterol retention phenotype conducive to foam cell formation. Therefore, only TG levels in large VLDL particles have no relationship with Coronary revascularization, while the other particles have a positive causal relationship with it.

We can look at Figure 3, we will discuss lines 1-10 and lines 15-26, the first part is lines 1-10, and the second part is lines 15-26. Compare the two, we can see two differences: the first is whether there is a positive causal relationship, and the second is the difference in trends. After replacing TG levels with TG to total lipids ratio, the causal relationship changed from positive to non-positive. The trend also changed from basically unchanged (the causal relationship in the second part changed little) to gradually decreasing (the causal relationship in the first part changed from negative to none). The higher ratio of TG to total lipids is conducive to the exchange (CETP transfers CE from HDL to VLDL in exchange for TG to HDL). The accumulation of exchange products prevents the exchange from continuing. this exchange also reduces this ratio (the ratio of TG to total lipids). These changes show up in a trend where negative causality decreases until it reaches zero. There are not a lot of products accumulation of this exchange (CETP transferring CE from HDL to VLDL in exchange for TG to HDL) in lines 1-5 of Figure 3, it is conducive to the play of the function of CETP. With the progress of metabolism, more and more products accumulate, which is not conducive to the play of the function of CETP. If the function of CETP is not hindered, there is a negative causal relationship (just as lines 1-5 in Figure 3). If its function is impeded, it becomes irrelevant (just as lines 6-10 in Figure 3).

If we replace the TG levels (lines15-26 of Figure 3) with the ratio of TG to total lipids (lines 1-10 of Figure 3) under the same conditions. The higher ratio of TG to total lipids in different sizes of VLDL and LDL includes the increase of TG and the decrease of total lipids, a reduction in cholesterol can also lead to a total lipids reduction, which is conducive to the play of the function of CETP. If its function is promoted, the anti-atherosclerotic effects are enhanced. There is a negative causal relationship (just as lines 1-5 in Figure 3), or no causal relationship (just as lines 6-10 in Figure 3). Overall, their anti-atherosclerotic effect is enhanced and their pro-atherogenic effect is weakened compared to before the replacement.

For very large HDL, large HDL, medium HDL particles, the opposite is true. Replacing the TG levels (lines27-29 of Figure 3) with the ratio of TG to total lipids (lines11-13 of Figure 3) under the same conditions, the result changed from no to positive causal relationship. The higher ratio of TG to total lipids includes an increase in TG and a reduction in cholesterol, which is not conducive to the play of the function of CETP. If its function is impeded, the pro-atherosclerotic effects are enhanced. This is demonstrated by comparing data from lines 27-29 with those in lines 11-13 of Figure 3. The causal relationship changed from no to positive. Overall, their pro-atherosclerotic effects are enhanced and their anti-atherogenic effects are weakened compared to before replacement.

Our study demonstrates that modulating CETP function - specifically its activation level - is a key determinant of anti-atherosclerotic efficacy. CETP facilitates cholesterol ester (CE) transfer from HDL to a broader spectrum of lipoproteins including CM, VLDL, IDL, and LDL. This mechanism enhances the reverse cholesterol transport by distributing cholesterol to diverse lipoprotein particles that can be processed by multiple metabolism-related receptors.

We found that different lipoprotein subclasses differentially influence CETP functionality and consequent anti-atherosclerotic effects. Specifically, the triglyceride-to-total lipid ratio in CM and VLDL subfractions (extremely large to small VLDL) promotes normal CETP function. This enhancement of cholesterol clearance capacity shows a negative causal association with coronary revascularization risk. In contrast, other lipoprotein particles examined in our study showed no such protective causal correlation.”

The ratio of TG to the total /TG levels in small HDL had positive causal relationship with Coronary revascularization, First, in humans HDL-FC has a plasma halftime of nearly 10 min[23] [24], too fast for meaningful esterification by LCAT and far shorter than the lifetime of HDL-proteins (∼6 days)[23][25]. Thus, spontaneous transfer is responsible for much of the total cholesterol biodistribution and cytotoxicity, particularly in the state of high plasma free cholesterol[23][26][27][28]. Indeed, cellular free cholesterol accumulation underlies foam cell death[23][29]. Collectively, these data support the hypothesis that most hepatic extraction of HDL-FC occurs without esterification. Not all HDL-FC is hepatically extracted. Free cholesterol transfers to nearly all other tissues, especially erythrocytes[23][28][30]. Free cholesterol transfer rates increase with decreasing HDL size [23], so the smaller HDL would have greater bioavailability[23]. Free cholesterol can diffuse from HDL, causing cytotoxicity, especially in small HDL. Which means that the pro-atherosclerotic effect of small HDL is minimally affected by the changes in lipid level and lipid ratio. This is demonstrated by comparing data from lines 14(TG to total lipids ratio in small HDL [P=4.19E-05, OR:1.44,95% CI:1.21-1.71]) with those in lines 30(TG levels in small HDL [P=3.66E-04, OR:1.44,95% CI:1.18-1.76]) of Figure 3. Which has little effect on the result after the interchange of TG levels and the ratio of TG to total lipids in small HDL. However, in very large HDL, large HDL, medium HDL, this exchange can have significant impacts on the results. Replacing the TG levels (lines27-29 of Figure 3) with the ratio of TG to total lipids (lines11-13 of Figure 3), the causal relationship changed from no to positive. Overall, their pro-atherosclerotic effects are enhanced and their anti-atherogenic effects are weakened compared to before the replacement.

Our findings demonstrate distinct atherogenic pathways mediated by HDL subclasses: while spontaneous free cholesterol (FC) transfer drives pro-atherosclerotic effects in small HDL particles, these pathological outcomes in large HDL particles primarily arise from mutual exchanges of lipid components.

There was no causal relationship between TG levels in (very large HDL, large HDL, medium HDL) and Coronary revascularization, but the ratio of TG to total lipids in (very large HDL, large HDL, medium HDL) had a positive causal correlation with Coronary revascularization. If HDL contains too much cholesterol. CETP transfers CE from HDL to VLDL in exchange for TG to HDL, hepatic lipase hydrolyzes TG in HDL, in turn promoting the reverse cholesterol transport. The rise TG to total lipids ratio is the result of excessive exchange (CETP transfers CE from HDL to VLDL in exchange for TG to HDL). It also prevents further exchange. The former condition produces more cholesterol-rich protein particles than the metabolism-related receptor can process. It has the effect of promoting atherosclerosis. The second condition reduces the ability to transport cholesterol lipids to other lipoprotein particles. It prevents the subsequent reverse transport of cholesterol and its uptake and utilization by cells. Both of them can increase the “non-metabolism-related pro-atherogenic pathway”, and reduce the “promote-cholesterol-metabolism-effect”. Our research reveals that TG to total lipids ratio in (very large HDL, large HDL, medium HDL) has a positive causal correlation with Coronary revascularization, this condition may be due to the compensatory response of the body caused by excessive cholesterol levels. However, it will hinder the subsequent reverse cholesterol transport. increase the deleterious effects of promoting atherosclerosis.

## Conclusions

The causal relationship between lipoproteins and coronary revascularization (ANGIO or CABG) shifts from negative to no, and then to positive, due to changes in the ratio of TG to total lipids within different lipoprotein particles.

This ratio variation across lipoprotein types and sizes differentially modulates CETP activity. When CETP transfers cholesterol esters (CE) from HDL to other lipoproteins — such as chylomicrons (CM), VLDL, IDL, and LDL — these particles become substrates for a broader range of metabolism-related receptors.

Lipoproteins vary in type, lipid level, and lipid ratio. There are different types of lipoprotein receptors that are expressed in various tissues and cells. The binding of Different lipoproteins to different receptors in various tissues and cells can produce different antiatherogenic and proatherogenic effects. The final result depends on their total effect, which results in a negative, positive, or no causal relationship between different lipoprotein subfractions and Coronary revascularization (ANGIO or CABG).

## Data Availability

The datasets generated during the current study are not publicly available, but are available from the corresponding author on reasonable request.

https://gwas.mrcieu.ac.uk/datasets/

## Acknowledgements

We are very grateful to the IEU Open GWAS project and the FinnGen. We are very grateful to these databases for providing data to our Mendelian randomization study. We are very grateful to the researchers who shared the data and the participants to provide great help for our Mendelian randomization study.

## Footnote

## Nonstandard Abbreviations and Acronyms

ANGIO: Angioplasty
ApoER2: apolipoprotein E receptor 2
AS: atherosclerosis
CABG: coronary artery bypass grafting
CE: cholesterol esters
CETP: Cholesteryl Ester Transfer Protein
CI: confidence interval
CM: chylomicrons
FC: free cholesterol
LCAT: Lecithin: Cholesterol Acyltransferase
LDLR: low-density lipoprotein receptor
MR: Mendelian Randomization
SNPs: single nucleotide polymorphisms
SR-B1: Class B Scavenger Receptor B1
TG: triglycerides
VLDL: Very Low-Density Lipoproteins
VLDLR: Very Low-Density Lipoproteins receptor

## References

[1] Feingold KR. Introduction to Lipids and Lipoproteins. [Updated 2024 Jan 14]. In: Feingold KR, Anawalt B, Blackman MR, et al., editors. Endotext [Internet]. South Dartmouth (MA): MDText.com, Inc.; 2000-. Available from: https://www.ncbi.nlm.nih.gov/books/NBK305896/

[2] Larsson SC, Butterworth AS, Burgess S. Mendelian randomization for cardiovascular diseases: principles and applications. Eur Heart J. 2023 Dec 14;44(47):4913–4924. 10.1093/eurheartj/ehad736. PMID: 37935836; PMCID: PMC10719501.

[3] Mineo C. Lipoprotein receptor signalling in atherosclerosis. Cardiovasc Res. 2020 Jun 1;116(7):1254–1274. 10.1093/cvr/cvz338. PMID: 31834409; PMCID: PMC7243280.

[4] Linton MF, Tao H, Linton EF, Yancey PG. SR-BI: A Multifunctional Receptor in Cholesterol Homeostasis and Atherosclerosis. Trends Endocrinol Metab. 2017 Jun;28(6):461–472. 10.1016/j.tem.2017.02.001. Epub 2017 Mar 1. PMID: 28259375; PMCID: PMC5438771.

[5] Silverstein RL, Febbraio M. CD36, a scavenger receptor involved in immunity, metabolism, angiogenesis, and behavior. Sci Signal. 2009 May 26;2(72):re3. 10.1126/scisignal.272re3. PMID: 19471024; PMCID: PMC2811062.

[6] Dieckmann M, Dietrich MF, Herz J. Lipoprotein receptors--an evolutionarily ancient multifunctional receptor family. Biol Chem. 2010 Nov;391(11):1341–63. 10.1515/BC.2010.129. PMID: 20868222; PMCID: PMC3529395.

[7] Krieger M, Herz J. Structures and functions of multiligand lipoprotein receptors: macrophage scavenger receptors and LDL receptor-related protein (LRP). Annu Rev Biochem. 1994;63:601–37. 10.1146/annurev.bi.63.070194.003125. PMID: 7979249.

[8] Kattoor AJ, Goel A, Mehta JL. LOX-1: Regulation, Signaling and Its Role in Atherosclerosis. Antioxidants (Basel). 2019 Jul 11;8(7):218. 10.3390/antiox8070218. PMID: 31336709; PMCID: PMC6680802.

[9] Go GW, Mani A. Low-density lipoprotein receptor (LDLR) family orchestrates cholesterol homeostasis. Yale J Biol Med. 2012 Mar;85(1):19–28. Epub 2012 Mar 29. https://pubmed.ncbi.nlm.nih.gov/22461740/PMID: 22461740; PMCID: PMC3313535.

[10] Willnow TE, Nykjaer A, Herz J. Lipoprotein receptors: new roles for ancient proteins. Nat Cell Biol. 1999 Oct;1(6):E157–62. 10.1038/14109. PMID: 10559979

[11] Li Y, Cam J, Bu G. Low-density lipoprotein receptor family: endocytosis and signal transduction. Mol Neurobiol. 2001 Feb;23(1):53–67. 10.1385/MN:23:1:53. PMID: 11642543.

[12] Ye ZJ, Go GW, Singh R, Liu W, Keramati AR, Mani A. LRP6 protein regulates low density lipoprotein (LDL) receptor-mediated LDL uptake. J Biol Chem. 2012 Jan 6;287(2):1335–44. 10.1074/jbc.M111.295287. Epub 2011 Nov 28. PMID: 22128165; PMCID: PMC3256876.

[13] Ishibashi S, Brown MS, Goldstein JL, Gerard RD, Hammer RE, Herz J. Hypercholesterolemia in low density lipoprotein receptor knockout mice and its reversal by adenovirus-mediated gene delivery. J Clin Invest. 1993 Aug;92(2):883–93. 10.1172/JCI116663. PMID: 8349823; PMCID: PMC294927.

[14] Yagyu H, Lutz EP, Kako Y, Marks S, Hu Y, Choi SY, Bensadoun A, Goldberg IJ. Very low density lipoprotein (VLDL) receptor-deficient mice have reduced lipoprotein lipase activity. Possible causes of hypertriglyceridemia and reduced body mass with VLDL receptor deficiency. J Biol Chem. 2002 Mar 22;277(12):10037–43. 10.1074/jbc.M109966200. Epub 2002 Jan 14. PMID: 11790777.

[15] Terrand J, Bruban V, Zhou L, Gong W, El Asmar Z, May P, Zurhove K, Haffner P, Philippe C, Woldt E, Matz RL, Gracia C, Metzger D, Auwerx J, Herz J, Boucher P. LRP1 controls intracellular cholesterol storage and fatty acid synthesis through modulation of Wnt signaling. J Biol Chem. 2009 Jan 2;284(1):381–388. 10.1074/jbc.M806538200. Epub 2008 Nov 6. PMID: 18990694; PMCID: PMC2610522.

[16] Cabezas F, Lagos J, C é spedes C, Vio CP, Bronfman M, Marzolo MP. Megalin/LRP2 expression is induced by peroxisome proliferator-activated receptor -alpha and -gamma: implications for PPARs’ roles in renal function. PLoS One. 2011 Feb 2;6(2):e16794. 10.1371/journal.pone.0016794. PMID: 21311715; PMCID: PMC3032793.

[17] Strickland DK, Au DT, Cunfer P, Muratoglu SC. Low-density lipoprotein receptor-related protein-1: role in the regulation of vascular integrity. Arterioscler Thromb Vasc Biol. 2014 Mar;34(3):487–98. 10.1161/ATVBAHA.113.301924. Epub 2014 Feb 6. PMID: 24504736; PMCID: PMC4304649.

[18] Yamada Y, Doi T, Hamakubo T, Kodama T. Scavenger receptor family proteins: roles for atherosclerosis, host defence and disorders of the central nervous system. Cell Mol Life Sci. 1998 Jul;54(7):628–40. 10.1007/s000180050191. PMID: 9711230; PMCID: PMC11147329.

[19] PrabhuDas MR, Baldwin CL, Bollyky PL, Bowdish DME, Drickamer K, Febbraio M, Herz J, Kobzik L, Krieger M, Loike J, McVicker B, Means TK, Moestrup SK, Post SR, Sawamura T, Silverstein S, Speth RC, Telfer JC, Thiele GM, Wang XY, Wright SD, El Khoury J. A Consensus Definitive Classification of Scavenger Receptors and Their Roles in Health and Disease. J Immunol. 2017 May 15;198(10):3775–3789. 10.4049/jimmunol.1700373. PMID: 28483986; PMCID: PMC5671342.

[20] Goldstein JL, Brown MS. A century of cholesterol and coronaries: from plaques to genes to statins. Cell. 2015 Mar 26;161(1):161–172. 10.1016/j.cell.2015.01.036. PMID: 25815993; PMCID: PMC4525717.

[21] Borrell-Pagès M, Romero JC, Badimon L. LRP5 deficiency down-regulates Wnt signalling and promotes aortic lipid infiltration in hypercholesterolaemic mice. J Cell Mol Med. 2015 Apr;19(4):770–7. 10.1111/jcmm.12396. Epub 2015 Feb 5. PMID: 25656427; PMCID: PMC4395191.

[22] Fujino T, Asaba H, Kang MJ, Ikeda Y, Sone H, Takada S, Kim DH, Ioka RX, Ono M, Tomoyori H, Okubo M, Murase T, Kamataki A, Yamamoto J, Magoori K, Takahashi S, Miyamoto Y, Oishi H, Nose M, Okazaki M, Usui S, Imaizumi K, Yanagisawa M, Sakai J, Yamamoto TT. Low-density lipoprotein receptor-related protein 5 (LRP5) is essential for normal cholesterol metabolism and glucose-induced insulin secretion. Proc Natl Acad Sci U S A. 2003 Jan 7;100(1):229–34. 10.1073/pnas.0133792100. Epub 2002 Dec 30. PMID: 12509515; PMCID: PMC140935.

[23] Gillard BK, Rosales C, Gotto AM Jr, Pownall HJ. The pathophysiology of excess plasma-free cholesterol. Curr Opin Lipidol. 2023 Dec 1;34(6):278–286. 10.1097/MOL.0000000000000899. Epub 2023 Sep 25. PMID: 37732779; PMCID: PMC10624414.

[24] Schwartz CC, VandenBroek JM, Cooper PS. Lipoprotein cholesteryl ester production, transfer, and output in vivo in humans. J Lipid Res. 2004 Sep;45(9):1594–607: 10.1194/jlr.M300511-JLR200. Epub 2004 May 16. PMID: 15145983.

[25] Blum CB, Levy RI, Eisenberg S, Hall M 3rd, Goebel RH, Berman M. High density lipoprotein metabolism in man. J Clin Invest. 1977 Oct;60(4):795–807. 10.1172/JCI108833. PMID: 197124; PMCID: PMC372427.

[26] Dole VS, Matuskova J, Vasile E, Yesilaltay A, Bergmeier W, Bernimoulin M, Wagner DD, Krieger M. Thrombocytopenia and platelet abnormalities in high-density lipoprotein receptor-deficient mice. Arterioscler Thromb Vasc Biol. 2008 Jun;28(6):1111–6. 10.1161/ATVBAHA.108.162347. Epub 2008 Apr 24. PMID: 18436807; PMCID: PMC2683374.

[27] Holm TM, Braun A, Trigatti BL, Brugnara C, Sakamoto M, Krieger M, Andrews NC. Failure of red blood cell maturation in mice with defects in the high-density lipoprotein receptor SR-BI. Blood. 2002 Mar 1;99(5):1817–24. 10.1182/blood.v99.5.1817. PMID: 11861300.

[28] Liu J, Gillard BK, Yelamanchili D, Gotto AM Jr, Rosales C, Pownall HJ. High Free Cholesterol Bioavailability Drives the Tissue Pathologies in Scarb1-/- Mice. Arterioscler Thromb Vasc Biol. 2021 Oct;41(10):e453–e467.10.1161/ATVBAHA.121.316535. Epub 2021 Aug 12. PMID: 34380332; PMCID: PMC8458258.

[29] Kellner-Weibel G, Jerome WG, Small DM, Warner GJ, Stoltenborg JK, Kearney MA, Corjay MH, Phillips MC, Rothblat GH. Effects of intracellular free cholesterol accumulation on macrophage viability: a model for foam cell death. Arterioscler Thromb Vasc Biol. 1998 Mar;18(3):423–31. 10.1161/01.atv.18.3.423. PMID: 9514411.

[30] Xu B, Gillard BK, Gotto AM Jr, Rosales C, Pownall HJ. ABCA1-Derived Nascent High-Density Lipoprotein-Apolipoprotein AI and Lipids Metabolically Segregate. Arterioscler Thromb Vasc Biol. 2017 Dec;37(12):2260–2270. 10.1161/ATVBAHA.117.310290. Epub 2017 Oct 26. PMID: 29074589; PMCID: PMC5831354.

[31] Lund-Katz S, Hammerschlag B, Phillips MC. Kinetics and mechanism of free cholesterol exchange between human serum high- and low-density lipoproteins. Biochemistry. 1982 Jun 8;21(12):2964–9. 10.1021/bi00541a025. PMID: 7104306.

